# Analytic Choices Shape Genomic Risk Estimates from Electronic Health Records: Coronary Heart Disease in eMERGE IV

**DOI:** 10.64898/2026.04.28.26352002

**Authors:** Jingheng H. Chen, Sarah A. Knerr, David L. Veenstra, Noura S. Abul-Husn, Sarah C. Hanks, Eimear E. Kenny, Nita A Limdi, Josh B. Cortopassi, David Crosslin, Gail P. Jarvik, Iftikhar J. Kullo

**Affiliations:** Institute of Public Health Genetics, University of Washington, Seattle, WA, USA; Department of Health System and Population Health, School of Public Health, University of Washington, Seattle, WA, USA; Department of Pharmacy, School of Pharmacy, University of Washington, Seattle, WA, USA; Institute for Genomic Health, Icahn School of Medicine at Mount Sinai, New York, NY, USA; 23andMe Research Institute, Palo Alto, CA, USA; Department of Neurology, Heersink School of Medicine, The University of Alabama at Birmingham, Birmingham, AL, USA; Division of Biomedical Informatics and Genomics, Tulane University School of Medicine, New Orleans, LA, USA; Division of Medical Genetics, School of Medicine, University of Washington, Seattle, WA; Division of Cardiovascular Medicine and the Mellowes Center for Genomic Sciences and Precision Medicine, Medical College of Wisconsin, Milwaukee, WI, USA

## Abstract

**Background:** Electronic health records (EHR) are an important data source for genomic studies, but challenges exist in ascertaining cases and observation start time. We used data derived from the Electronic Medical Records and Genomics (eMERGE) IV study to examine how analytic assumptions regarding case ascertainment and EHR entry time influence estimation of monogenic and polygenic risks for coronary heart disease (CHD).

**Methods:** We assessed agreement between CHD cases ascertained from EHR phenotyping and survey. Associations of monogenic variants and high (top 5%) PRS of CHD were evaluated using multivariate relative risk (RR) regression under three alternative case definitions: EHR-algorithm-defined, self-reported, and combined. Time-to-event analyses (Kaplan–Meier method and Cox proportional hazards models) were conducted under three entry time specifications: (1) entry at the first EHR record, (2) entry at the start of the latest consecutive observation period, and (3) no left truncation.

**Results:** The agreement between CHD cases ascertained by the EHR-based algorithm versus self-report was 37.2% among individuals identified as cases by at least one source, with the EHR algorithm demonstrating higher accuracy. The adjusted RR [95% confidence interval (CI)] associated with high PRS was 2.05 [1.50, 2.81] for EHR-defined, 1.49 [1.04, 2.13] for self-reported, and 1.66 [1.27, 2.18] for combined CHD. Estimated cumulative incidence by age 75 was 0.188 using the first EHR code as left truncation and 0.225 using the most recent observation period. Hazard ratio (HR) estimates were similar across the three left truncation scenarios.

**Conclusion:** The choice of case definition meaningfully influenced RR estimates, whereas alternative specifications of EHR entry time affected absolute cumulative incidence estimates but has minimal impact on HR. These findings highlight the impact of analytical choices in EHR and survey-data-based studies that are applicable beyond the context of CHD.

## INTRODUCTION

Electronic health records (EHR) capture rich, real-world clinical information at scale and are increasingly used as a data source for epidemiological research.^1^ Many large cohort studies, including the UK Biobank^2^ and All of Us^3^, have linked EHR data to genomic data, enabling population-level studies of genetic associations with a range of conditions.

However, since EHR are designed for clinical documentation and billing rather than for research, repurposing EHR data for epidemiological studies introduces many analytical challenges.^4^ Two issues in particular threaten the internal validity of genetic association studies. First, phenotypes are commonly derived from diagnosis and procedure codes,^5^ which are susceptible to misclassification due to possible upcoding practices, documentation errors, or incomplete data capture resulting from fragmented healthcare utilization.^6,7^ Misclassification of case status can reduce statistical power and bias association estimates. Participant self-reported health history has been proposed as a preferred data source to improve case ascertainment;^8^ however, the relative accuracy of EHR-data-defined and self-reported cases remains uncertain, and no consensus exists regarding the optimal strategy for defining cases.^9–11^

Second, for analyses involving time-at-disease-onset, additional complications arise because individuals are typically not observed continuously from the time origin, which, in the context of germline genomic risk factors, is usually birth. This introduces a “delayed entry” (i.e., left truncation) bias that will inflate time-at-onset estimates if not corrected, typically via risk set adjustment whereby individuals are counted toward the risk set only after their prespecified observation entry time.^12,13^ In retrospective EHR-based studies, the entry time is often operationalized as the time of the earliest record in the EHR.^14^ However, early EHR data may be highly fragmented and not reflect the start of consistent observation; using these as entry times could inflate person-time observed and potentially introduce bias.

We sought to further investigate these two issues using the extensive EHR and survey data available from the Electronic Medical Records and Genomics (eMERGE) IV study^15^, in an analysis of monogenic and polygenic risks for coronary heart disease (CHD). CHD provides an informative exemplar because it is commonly ascertained using EHR billing codes and has age-dependent onset necessitating time-to-event modeling. Further, the associations of monogenic risk variants and polygenic risk scores (PRS) with CHD have been previously reported in multiple large EHR-based cohorts, with established implications in disease prevention.^16–21^ However, studies have employed heterogeneous case definitions and modeling choices, limiting their comparability.

In this study, we compared EHR-based and self-reported CHD case ascertainment in the eMERGE IV cohort and evaluated how the choice of case definition influenced relative risk estimates for monogenic and polygenic risk of CHD. We also investigated how alternative definitions of EHR entry time influenced age-specific cumulative risk and hazard ratio estimates. By quantifying the sensitivity of risk estimations to analytical assumptions regarding EHR observation time and outcomes, our work contributes to methodological considerations relevant to EHR-based genetic risk association studies beyond eMERGE IV and CHD.

## METHODS

### Population and Data Source

We used interim research data derived from the eMERGE IV study. The complete protocol of eMERGE IV has been published elsewhere.^15,22,23^ In brief, the study enrolled a cohort of ∼25000 pediatric and adult participants across 10 clinical sites in the US with the primary goal of assessing the healthcare impact of returning an integrated genome-informed risk assessment (GIRA) for 11 common complex diseases,^22^ including CHD for adult participants. Upon enrollment, study participants answered surveys regarding their demographics, health behaviors, and personal health history relevant to the conditions under study, provided biospecimens for genotyping and monogenic testing, and granted linkage to their EHR at their enrolling clinical site. Structured EHR data elements, including all ICD (International Classification of Diseases) and CPT (Current Procedural Terminology) codes, selected medication and laboratory measurements, along with the age-at-record, were extracted from the clinical sites for participants enrolled before August 2023 as part of interim quality assurance.

This analysis included participants who were adults (age ≥18 years) at enrollment, had completed all study-related activities with GIRA returned, were included in the August 2023 interim EHR data pull, and were not actively withdrawn as of April 2025 (n =12,353). We excluded participants missing either monogenic or PRS results, as these were the exposures of interest, and participants with less than 1 year of EHR data to ensure sufficient observation time. We additionally excluded one participant with CHD before age 17, as atherosclerotic CHD is rare among children and upon inspection the case likely reflected congenital coronary disease, a phenotype outside the scope of this analysis. The final analytical cohort contained 11,699 participants.

### CHD Case Ascertainment

CHD cases from structured EHR data were identified using a phenotyping algorithm developed in past iterations of eMERGE.^11,15^ The algorithm focuses on “hard” CHD cases, including myocardial infarction (MI) and cases that require coronary revascularization, including coronary artery bypass surgery (CABG) and percutaneous coronary intervention (PCI). Individuals with MI were defined by having at least two ICD-9 or 10 diagnostic codes for MI within a 5-day window. Individuals who underwent CABG or PCI were defined as those who had at least one relevant procedural (ICD-9 or ICD-10-PCS [Procedure Coding System], or CPT-4) code. A complete list of codes used can be found in **Supplemental Table 1**. Age at CHD onset was defined as the earliest of the first recorded MI (first out of the >2 codes), CABG, or PCI event.

Self-reported CHD status was determined based on participants’ answers to the following questions in the baseline survey: “*coronary heart disease: I currently have this condition*” and “*coronary heart disease: I had this condition in the past*”. Participants who answered positively to either question were classified as having self-reported CHD. For these participants, additional questions were asked about whether the participant had “*been hospitalized for a heart attack or myocardial infarction*”, had “*heart bypass surgery*”, or “*stenting of the heart arteries*”.

To assess agreement of case ascertainment, self-reported CHD status was compared to EHR algorithm-defined CHD status, and sub-phenotypes were matched to their closest EHR-algorithm defined counterparts (i.e., “heart attack” to MI, “heart bypass surgery” to CABG, and ‘‘stenting of the heart arteries” to PCI). For participants who had discrepant algorithm-defined and self-reported CHD status, we conducted a manual review of their EHR to determine the “ground truth” at a single clinical site (University of Washington Medicine). The review included 10 participants (randomly sampled out of 36) who reported having CHD on the survey but were classified as non-cases by the phenotyping algorithm (EHR algo-, self-report+), and 5 participants (exhaustive sample) who were classified as CHD positive by the algorithm but self-reported no CHD (EHR algo+, self-report-). Two reviewers examined all available EHRs for these participants and determined their CHD status based on relevant medical history.

We additionally defined a combined case status that classified a participant as a case if CHD was identified by either source for follow-up analysis.

### EHR Observation Periods and Entry Time Definitions

During exploratory analyses, we observed that early EHR data might contain artifacts that were unlikely to reflect actual clinical encounters (e.g., records of ICD-10 codes dated before the implementation of ICD-10). These spurious records occurred mostly during early ages of older participants and were followed by prolonged gaps in EHR data (e.g., participant ID_9 in **Figure 1A**). We reasoned that these could be backfilled artifacts and do not represent the start of consistent observation in the EHR.

**Figure 1.**
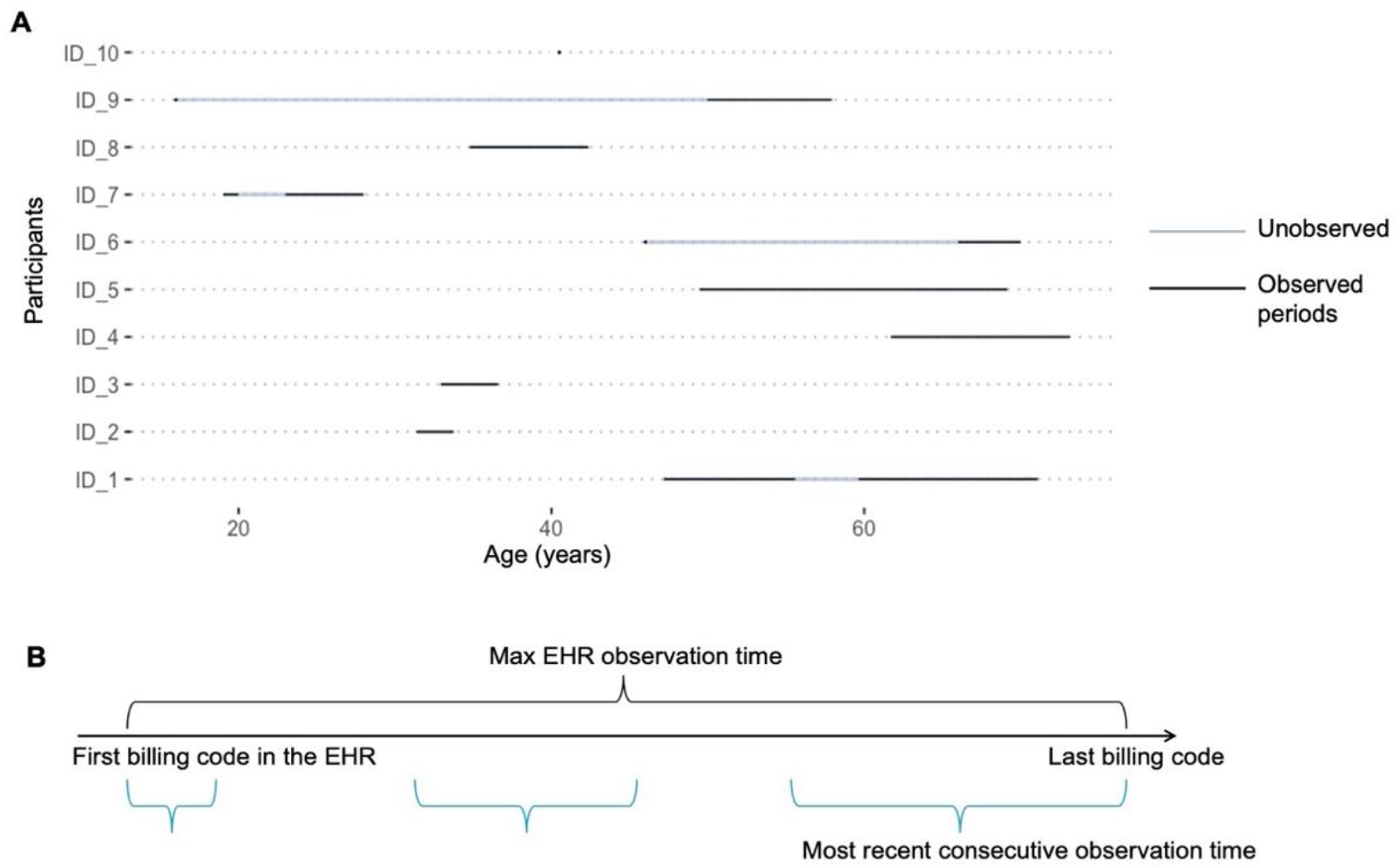
Illustration of inconsistent EHR observation time. **A)** Observed and unobserved periods for 10 randomly selected participants in the EHR cohort. Each horizontal line represents the participant’s age-based timeline under EHR observation, which starts at the age of the first recorded billing code. Shaded segments represent periods under continued observation, allowing gaps <1.5 years between codes. **B**) Schematic depiction of episodic EHR observation periods. Maximum EHR observation time is defined by the first and last billing code in one’s EHR. Consecutive observation periods are defined as spans of time with billing codes separated by gaps no greater than a prespecified threshold (1.5 years).

To investigate the continuity of EHR data capture, we calculated the following observation periods for each participant: (1) maximum EHR observation time, defined as time from the first billing code to the last billing code, and (2) consecutive observation periods, defined as time spans with billing codes occurring with gaps of less than 1.5 years (**Figure 1B**). The 1.5-year threshold was based on the Observational Medical Outcomes Partnership (OMOP) Common Data Model’s suggestion in determining EHR observation periods^25^. We compared the maximum and consecutive observation times to assess the length of observation gaps. The *start* of the *most recent* consecutive observation period was defined as an alternative EHR entry time for left truncation adjustment.

### Genomic Risk Factors

The exposures in this study were monogenic risk variants and high PRS for CHD. Monogenic risk for CHD is defined as having at least one copy of a pathogenic/likely pathogenic (P/LP)^26^ variant in *APOB, LDLR*, and *PCSK9*, or homozygous for a P/LP variant in *LDLRAP1*.^15^ The PRS used in the study was a multi-ancestry PRS model constructed using 458,384 single-nucleotide variants with summary statistics derived from a large multi-ancestry GWAS.^27,28^ Raw scores were standardized using the expected means and standard deviations based on the principal components of ancestry.^29^ A top 5 percentile cut-off was selected by eMERGE IV study workgroups as the “high-risk” threshold for CHD.^15^

### Statistical Analysis

#### Binary CHD outcomes

We described the proportion of participants with CHD ascertained using the EHR-based algorithm and survey, overall and by sex and age groups. The CHD risks associated with high PRS and monogenic risk variants were assessed using relative risk (RR) regression, implemented as modified Poisson regression with robust standard errors, the preferred approach for estimating RRs.^30^ We performed the same set of regression models for three outcome definitions: EHR-algorithm defined CHD, self-reported CHD, and the combined case status incorporating both sources. The exposures, monogenic risk and high PRS, were coded as two separate indicator variables with no interaction term, assuming additive effects. All associations were adjusted for the following covariates unless otherwise specified: age at enrollment, sex assigned at birth (hereafter, “sex”), clinical site, and maximum EHR observation length. Categorical covariates (sex, clinical site) were modeled using dummy variables, and continuous covariates (age, EHR observation length) were modeled linearly to the first order. Genetic principal components were not included because the resulted PRS had been ancestry-calibrated and normalized.^29^ Models were first fit in the entire cohort and then stratified by sex and 10-year age groups (<40, 40-49, 50-59, 60-69, >70 years).

#### Time-to-event analysis

All time-to-event analyses used EHR algorithm-defined CHD as the outcome, as self-report data did not include age at diagnosis. We used the Kaplan-Meier method to estimate the cumulative incidence of CHD by age, using age (years) as the time scale and right-censored at the age of the last billing code in the EHR. We tested three alternative left truncation scenarios: (1) first billing code as entry, (2) start of the most recent consecutive observation period as entry, (3) no left truncation (i.e., birth as entry) as control. For participants whose earliest billing codes included CHD-related codes, suggesting prevalent disease before entering the health system, the recorded age of the CHD event was retained. Confidence intervals for the cumulative incidence functions were constructed using Greenwood’s formula and complementary log-log transformation.^31^ We used Cox proportional hazards (PH) regression to assess the CHD hazard associated with high PRS and monogenic variants, separately under the three left truncation scenarios. Models used the same specification besides entry time. We adjusted for sex as a covariate and clinical site through stratification to relax the PH assumption among sites.

All analyses were performed in R (version 4.4.2). Time-to-event analyses were performed using the survival and survminer packages.^32,33^ Hypothesis tests for individual parameters were performed using two-sided Wald tests with α of 0.05.

## RESULTS

### Participants Characteristics

The study cohort consisted of 11,699 participants (mean age at enrollment 51.2 ±14.9 years, 67.4% female), recruited from 10 clinical sites. The median maximum EHR observation length was 10.51 years ([interquartile range (IQR)] 6.32-15.54 years). The breakdown of participant characteristics, including enrolled site, is shown in **Supplemental Table 2**.

### Comparison of EHR-derived and Self-reported Case Ascertainment

The EHR data-based phenotyping algorithm identified 455 participants (3.9%) with CHD, and 500 participants (4.4%) reported CHD on the baseline survey (**Table 1, left panel**). Among the 11,336 participants with both data sources available, overall agreement was 96.2%; however, this was driven largely by the 93.9% identified as CHD-negative by both sources. When only considering the 689 participants classified as CHD-positive by at least one source, agreement decreased to 37.2%, reflecting limited overlap between algorithm-defined and self-reported cases (**Table 1, right panel**). Out of the three sub-phenotypes, MI/ heart attack had the highest disagreement rate – 69 out of 178 (38.8%) participants self-reported having been “hospitalized for heart attack” were not identified as having MI using the EHR algorithm, and 16 out of 125 (12.6%) vice versa.

**Table 1.**
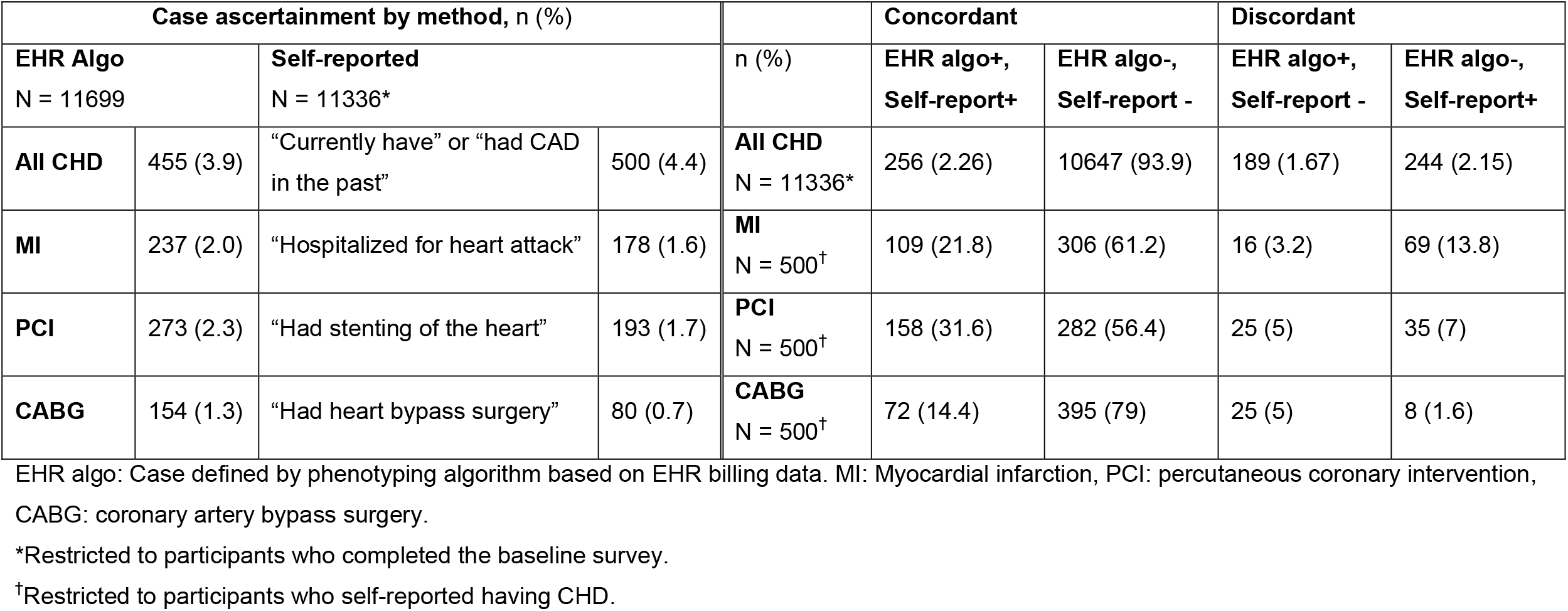
Case ascertainment from EHR-data based phenotyping algorithm and self-report.

Through manual chart review for the 15 discordant cases (see methods for selection criteria), we found definitive evidence of CHD for 2 out of 10 individuals who were EHR algo-, self-report+, and 3 out of 5 individuals who were EHR algo+, self-report-(**Supplemental Table 3**). In summary, among the 15 cases reviewed, the EHR algorithm was accurate for 11 (73%), compared to 4 (27%) cases by self-report. Six (out of 8) participants who self-reported CHD but were deemed not to have CHD by manual review had other cardiac conditions documented in the clinical notes (e.g., arrhythmia), but did not have evidence of CHD. Of the two participants who were classified as CHD by EHR algorithm but deemed not to have CHD by chart review, one had a similar procedure (e.g., stenting of arteries other than coronary) and the other had discussions of CHD risk with the clinician because of family history. Two self-reported cases not identified by the EHR algorithm received cardiac care outside of the health system.

### Relative Risk Estimates with Alternative Case Definitions

The relative risk (RR) estimates for monogenic risk and high (top 5%) PRS showed similar trends across analyses using EHR algorithm-defined, self-reported, or combined CHD outcomes, although some effect size magnitudes and precision varied, leading to differences in statistical inference (**Table 2**). For example, in the overall analysis, the adjusted RR associated with high PRS was 2.05 (95% CI: [1.50, 2.81], p<0.001) for EHR-defined CHD, but was 1.49 (95% CI: [1.04, 2.13], p<0.05) for self-reported and 1.66 (95% CI: [1.27, 2.18], p<0.001) for combined CHD. The adjusted RR associated with monogenic risk was 2.00 (95% CI: [1.01, 3.92], p<0.05) for self-reported CHD, but was lower and statistically non-significant for EHR-defined or combined CHD.

**Table 2.**
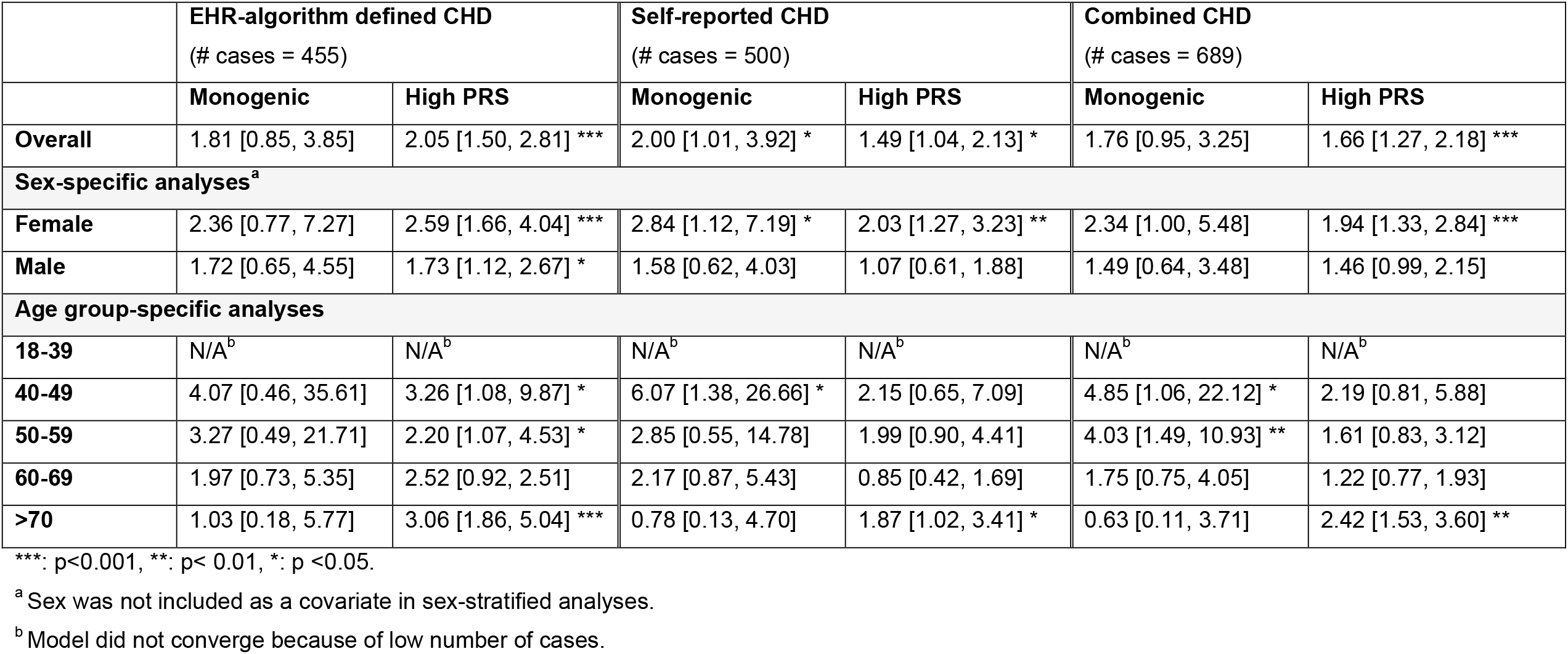
Relative risk of CHD for monogenic and polygenic risk factors, under three CHD definitions, RR [95% CI].

Despite some variability, the RR point estimates were generally larger among females and the middle-aged (40-49 and 50-59 yrs) groups for both risk factors across the models. Monogenic risk did not reach statistical significance in any stratum for EHR-defined CHD, but models using self-reported CHD yield an RR of 2.84 (95% CI: [1.12, 7.19], p<0.05) in the female-only stratum and 6.07 (95% CI: [1.38, 26.66], p<0.05) in the age 40-49 stratum.

### Variability in EHR Entry Time

The maximum EHR observation time equaled the most recent consecutive observation period for 52.8% of the participants. However, there were 3.76% of participants with substantial (>20 years) discrepancies between their maximum and most recent consecutive observation period, with the highest discrepancy being 59.9 years (**Figure 2A**). When EHR entry was defined by the start of the most recent consecutive period, the distribution of age-at-entry shifted toward older age (median shifted from 40.18 to 45.02 years), compared to by the first billing code in the EHR (**Figure 2B**).

**Figure 2.**
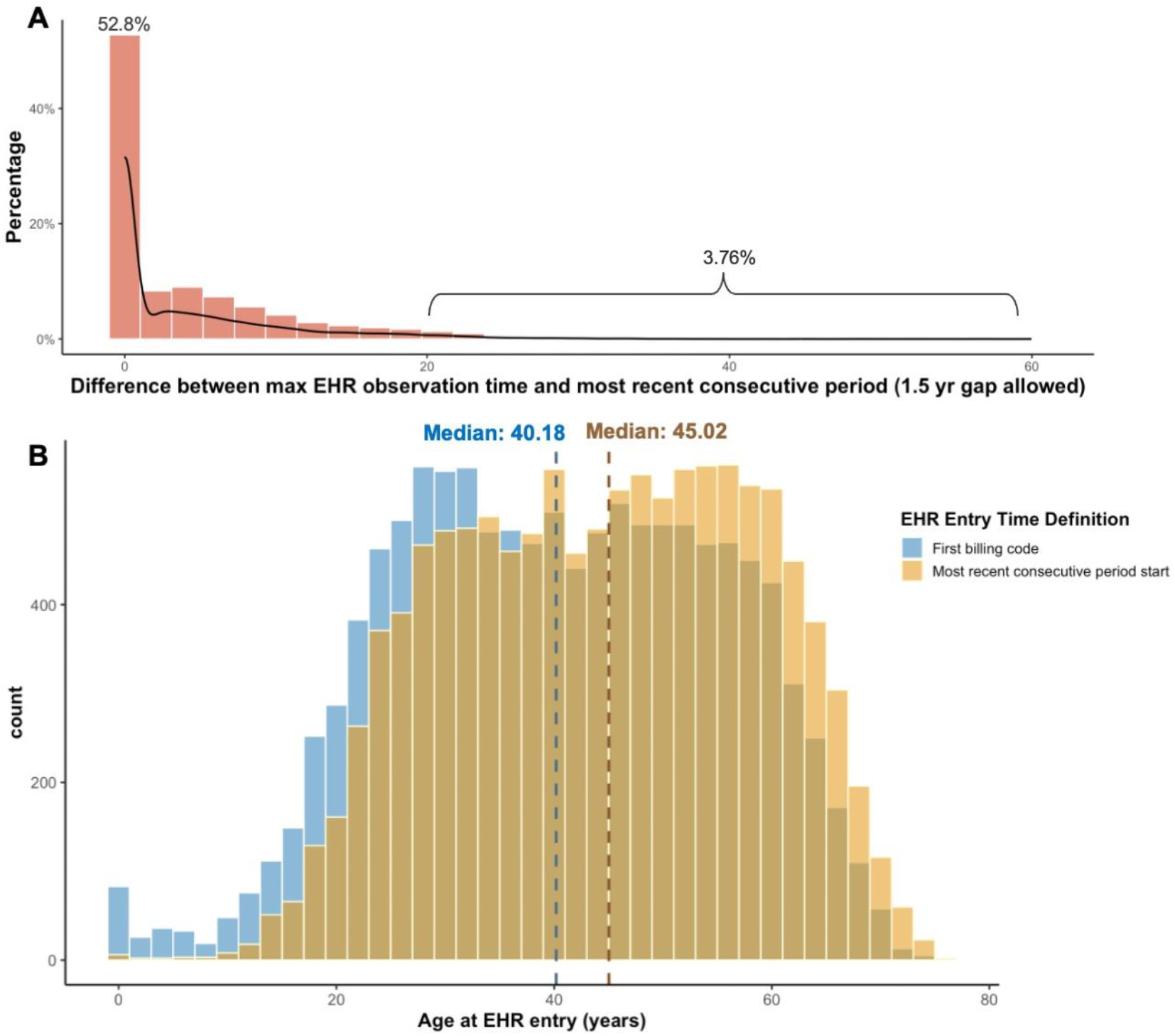
**A)** Difference between the maximum EHR observation time and the most recent consecutive observation period. **B)** Distribution of age at EHR entry by two definitions.

### Sensitivity of Risk Estimates to EHR Entry Time

Using the start of the most recent consecutive period as entry time yielded higher age-specific Kaplan-Meier cumulative incidence estimates than using the first EHR code as entry time, and both approaches yielded higher cumulative incidence estimates than the no left truncation scenario (**Figure 3**). The cumulative incidence by age 75 was estimated to be 0.149 (95% CI: [0.131, 0.169]) when left truncation was not adjusted, compared to 0.188 (95% CI: [0.168, 0.209]) when using the first EHR code as entry time and 0.225 (95% CI: [0.202, 0.249]) when using the start of most recent consecutive period as entry time (**Supplemental Table 4**).

**Figure 3.**
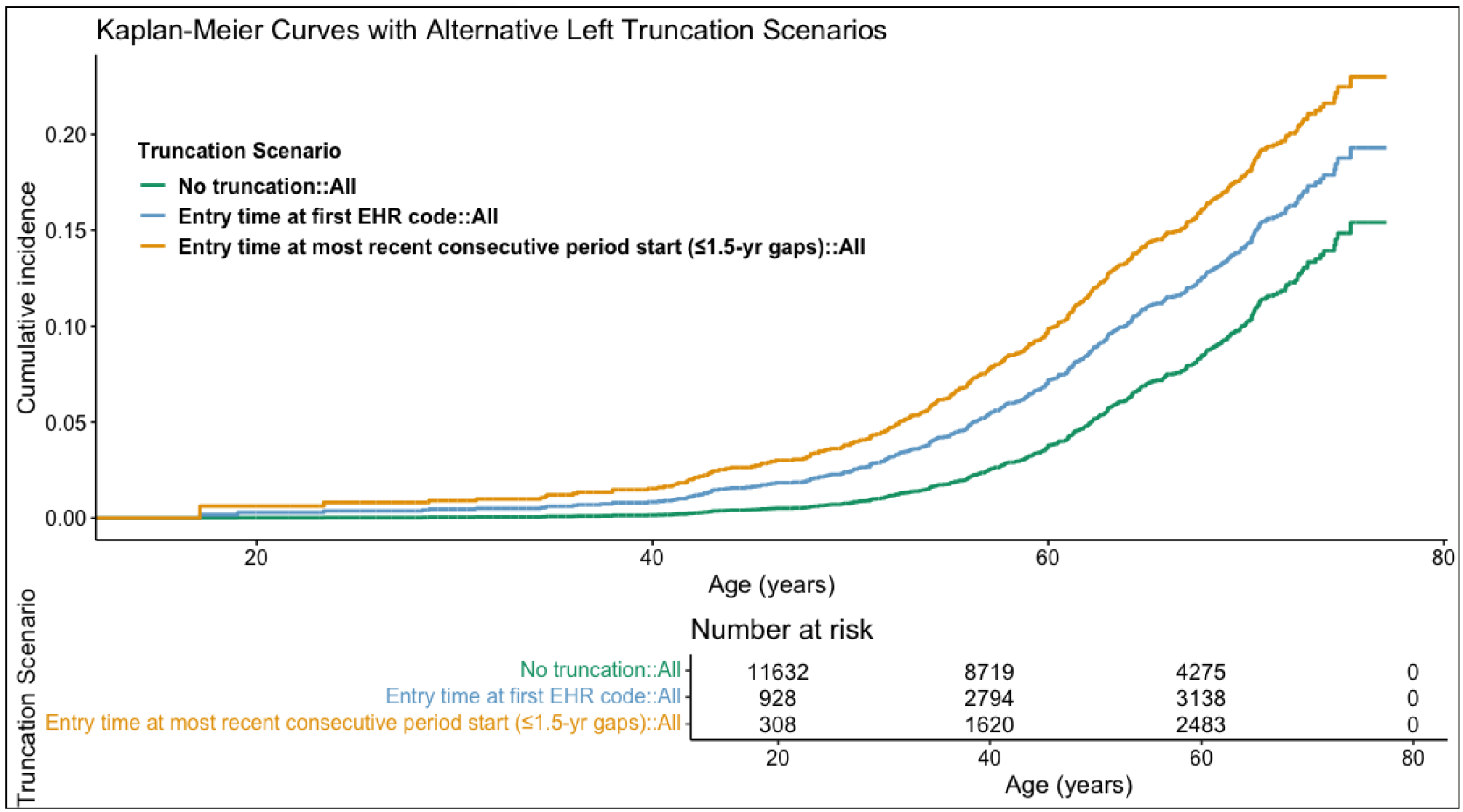
Kaplan-Meier curves for cumulative incidence of EHR algorithm-defined CHD, under three alternative left truncation scenarios. The numbers of participants in the risk set at age 20, 40, 60, and 80 across different left truncation scenarios are displayed in the table in the lower panel.

The hazard ratio (HR) estimates for monogenic risk and high (top 5%) PRS were similar across the three left truncation scenarios in the overall, sex-stratified, and most age groups in the age-stratified analyses (**Table 3**). In the overall model, the HR for CHD associated with high PRS ranged from 2.13 (95% CI: [1.52, 2.99], p<0.001) using the most recent consecutive period as entry time, to 2.24 (95% CI: [1.61, 3.13], p<0.001), using the first billing code as entry time. The HR associated with monogenic variants ranged from 1.84 (95% CI: [0.87, 3.90]), using the most consecutive period as entry time, to 1.86 (95% CI: [0.88, 3.93]), under the no truncation scenario. Similar to the RR models, the HR point estimates were more prominent among females and middle-aged groups. The HR estimate for monogenic risk in the 40-49 age group had the most variability, likely due to the small number of events in this group.

**Table 3.** Hazard ratio of monogenic and polygenic risk factors under three left truncation scenarios, HR [95% CI].

|  | No truncation |  | First billing code as entry time |  | Most recent consecutive period start as entry time |  |
| --- | --- | --- | --- | --- | --- | --- |
|  | Monogenic | High PRS | Monogenic | High PRS | Monogenic | High PRS |
| **Overall** | 1.86 [0.88, 3.93] | 2.17 [1.56, 3.03] *** | 1.85 [0.88, 3.92] | 2.24 [1.61, 3.13] *** | 1.84 [0.87, 3.90] | 2.13 [1.52, 2.99] *** |
| **Sex-specific analyses<sup>a</sup>** |  |  |  |  |  |  |
| **Female** | 2.19 [0.70, 6.88] | 2.73 [1.71, 4.34] *** | 2.56 [0.81, 8.05] | 2.59 [1.63, 4.12] *** | 2.42 [0.77, 7.63] | 2.65 [1.66, 4.22] *** |
| **Male** | 1.85 [0.69, 5.00] | 1.87 [1.15, 3.03] * | 1.75 [0.65, 4.74] | 1.75 [0.65, 4.74] ** | 1.83 [0.68, 4.97] | 1.77 [1.07, 2.92] * |
| **Age group-specific analyses** |  |  |  |  |  |  |
| **18-39** | N/A<sup>b</sup> | N/A<sup>b</sup> | N/A<sup>b</sup> | N/A<sup>b</sup> | N/A<sup>b</sup> | N/A<sup>b</sup> |
| **40-49** | 4.29 [0.56, 32.71] | 3.11 [0.92, 10.55] | 3.74 [0.49, 28.58] | 3.16 [0.93, 10.73] | 6.52 [0.80, 52.91] | 3.60 [1.05, 12.39] * |
| **50-59** | 3.48 [0.48, 25.50] | 2.39 [1.09, 5.24] * | 3.61 [0.49, 26.61] | 2.37 [1.08, 5.19] * | 3.92 [0.53, 29.14] | 2.71 [1.23, 5.97] * |
| **60-69** | 1.98 [0.73, 5.35] | 1.50 [0.88, 2.55] | 1.83 [0.68, 4.95] | 1.52 [0.90, 2.58] | 1.63 [0.60, 4.41] | 1.37 [0.79, 2.36] |
| **>70** | 1.28 [0.18, 9.20] | 3.49 [1.96, 6.24] *** | 1.35 [0.19, 9.78] | 4.08 [2.27, 7.30] *** | 1.30 [0.18, 9.43] | 3.54 [1.96, 6.42] *** |

\*\*\*: $p < 0.001$ , \*\*: $p < 0.01$ , \*: $p < 0.05$ .
<sup>a</sup> Sex was not included as a covariate in sex-stratified analyses.
<sup>b</sup> Model did not converge because of low number of cases.

## DISCUSSION

In this study, we investigated the impact of case ascertainment and EHR entry time on estimation of monogenic and polygenic risk associated with CHD in the eMERGE IV cohort. We observed moderate to low agreement between cases ascertained by the EHR-based algorithm and self-report and demonstrated that the choice of case definition meaningfully influenced effect estimation and statistical inference. In the time-to-event analyses considering alternative left truncation scenarios, we found that using the start of the most recent consecutive period as entry expectedly resulted in higher cumulative incidence estimates due to the smaller risk sets at earlier ages, but hazard ratio estimates remained consistent. Together, these findings highlight the impact of methodological choices in EHR-based studies that are applicable beyond the context of CHD.

Our findings on case ascertainment add to a growing body of literature that reported disagreement between EHR-derived and self-reported cases and underscore that neither source represents a definitive measure of disease status.^9–11^ Misclassification of cases by the EHR-based algorithm and self-report appears to arise from different mechanisms, which are non-random and, as a recent study suggests, can have substantial impacts on the downstream effect estimates and statistical power to detect association.^34^ Previous studies have also found that the relative performance of self-report and EHR-based case ascertainment varies by conditions and health systems,^10^ highlighting the need to use validated portable EHR algorithms and assess the context-specific performance of case definition when possible. Our validation results show that, in the context of atherosclerotic CHD within the eMERGE IV study, the algorithm based on EHR billing codes is more accurate than self-reported health history collected by survey and should be used for primary case definition.

Our analysis of EHR observation time highlights the fragmentation and potential artifactual errors in EHR data. Our approach of truncating early observations outside of the most recent consecutive period could help mitigate the artificial inflation of person-time observed caused by potentially spurious records; however, it might also exclude valid earlier clinical events and lead to misclassification of case status or time. This scenario mimics real-world data loss due to delayed entry to a health system and fragmented healthcare utilization. Although the cumulative incidence estimates were sensitive to alternative entry specifications, estimation of absolute age-specific risk is usually not the objective of EHR-based studies. The stability of hazard ratio estimates suggests that Cox models are not overly sensitive to the choice of EHR entry time. This robustness may generalize to settings with limited EHR data capture at earlier ages, alleviating previous concerns over bias from incomplete early EHR observations. Nonetheless, additional work, such as simulation studies, is needed to further clarify this question.

To our knowledge, the impact of EHR fragmentation on left truncation has not been systematically investigated before. Our study complements the literature on other analytical issues concerning the time-to-event models, including Korn et al. (1997) which recommended using age instead of time-on-study as the time scale in longitudinal studies^35^ and Irlmeier (2022) which found the Cox model is robust to delayed event time recorded in the EHR.^36^

Lastly, the estimated CHD risks associated with monogenic and PRS from our analyses are consistent with prior reports^16,18,27^, which collectively show that individuals with PRS at the upper tail of the distribution may have CHD risk at least comparable to those with high penetrance monogenic variants. Although our study has a methodological focus, our results also validated the associations of a multi-ancestry PRS with CHD in a new, independent cohort.

### Limitations

This study has several limitations. First, our study had a relatively small sample size, limiting our ability to produce precise estimates, especially for monogenic risk. Second, manual chart review was conducted on a limited, purposively selected subset of participants with discordant case classifications and was restricted to a single health system. As a result, the reviewed sample was not representative and could not be used to estimate sensitivity or specificity of the two CHD ascertainment methods. Third, we evaluated only one EHR-based phenotyping algorithm for CHD. The algorithm we used is more restrictive compared to phenotyping methods using a single occurrence of billing codes,^37,38^ which may generate different results. Fourth, our age-at-event analysis did not explicitly address left censoring or misclassification of event timing. For participants whose earliest recorded encounters included CHD-related billing codes, these diagnoses may represent prevalent disease occurring prior to entry into the health system. Failure to fully account for such left censoring could affect the results. Future studies should also assess whether case ascertainment accuracy and EHR data fragmentation patterns differ by social determinants of health.

### Conclusion

The choice of CHD case definition meaningfully influenced the estimated associations in relative risk regression models, while specifications of EHR entry time affected estimates of cumulative incidence, but not hazard ratios. These analytical decisions inherent to EHR-based research can shape study results and should be explicitly considered in the design and interpretation of survey and EHR-based genomic studies.

## Data Availability

The data used in this analysis are interim research data. Available upon request.

## ACKNOWLEDGEMENTS

We thank the participants and staff of the eMERGE IV study for their contributions. We thank Dr. Jessica Chubak, Dr. Elisabeth Rosenthal, Dr. Li Hsu for thoughtful discussions and feedback on data analysis, Dr. Mohammadreza Naderian for clinical guidance and Min Seon Park for assisting with manual chart review.

## SUPPLEMENTAL MATERIAL

**Supplemental Table 1. Billing codes used in the phenotyping algorithm See attachment**

**Supplemental Table 2.** Basic characteristics of participants in the study cohort.

|  | Overall cohort (N = 11699) |
| --- | --- |
| **Age at enrollment, yrs (mean± SD)** | 51.21 ±14.92 |
| **Age groups in years, n (%)** |  |
| 18-39 | 3065 (26.2) |
| 40-49 | 2050 (17.5) |
| 50-59 | 2355 (20.1) |
| 60-69 | 2886 (24.7) |
| >70 | 1343 (11.5) |
| **Sex at assigned birth, n (%)** |  |
| Female | 7889 (67.4) |
| Male | 3810 (32.6) |
| **Race and ethnicity, n (%)** |  |
| Asian | 967 (8.3) |
| Black, African American or African | 1843 (15.8) |
| Hispanic, Latino or Spanish | 1960 (16.8) |
| White | 5999 (51.3) |
| Other/ Admixed/ Prefer not to answer | 930 (8.0) |
| **Site, n (%)** |  |
| Vanderbilt | 680 (5.8) |
| UAB | 2466 (21.1) |
| Columbia | 1178 (10.1) |
| Mayo Clinic | 1063 (9.1) |
| MPHC (part of Mayo) | 278 (2.4) |
| UW | 1942 (16.6) |
| Northwestern | 1572 (13.4) |
| MGB | 1025 (8.8) |
| Mount Sinai | 1401 (12.0) |
| Cincinnati | 42 (0.4) |
| CHOP | 52 (0.4) |
| **High risk PRS, n (%)** | 515 (4.4) |
| **Monogenic risk, n (%)** | 83 (0.7) |
| *APOB* | 14 (0.1) |
| *LDLR* | 65 (0.6) |
| *PCSK9* | 4 (0.0) |

**Supplemental Table 3.**
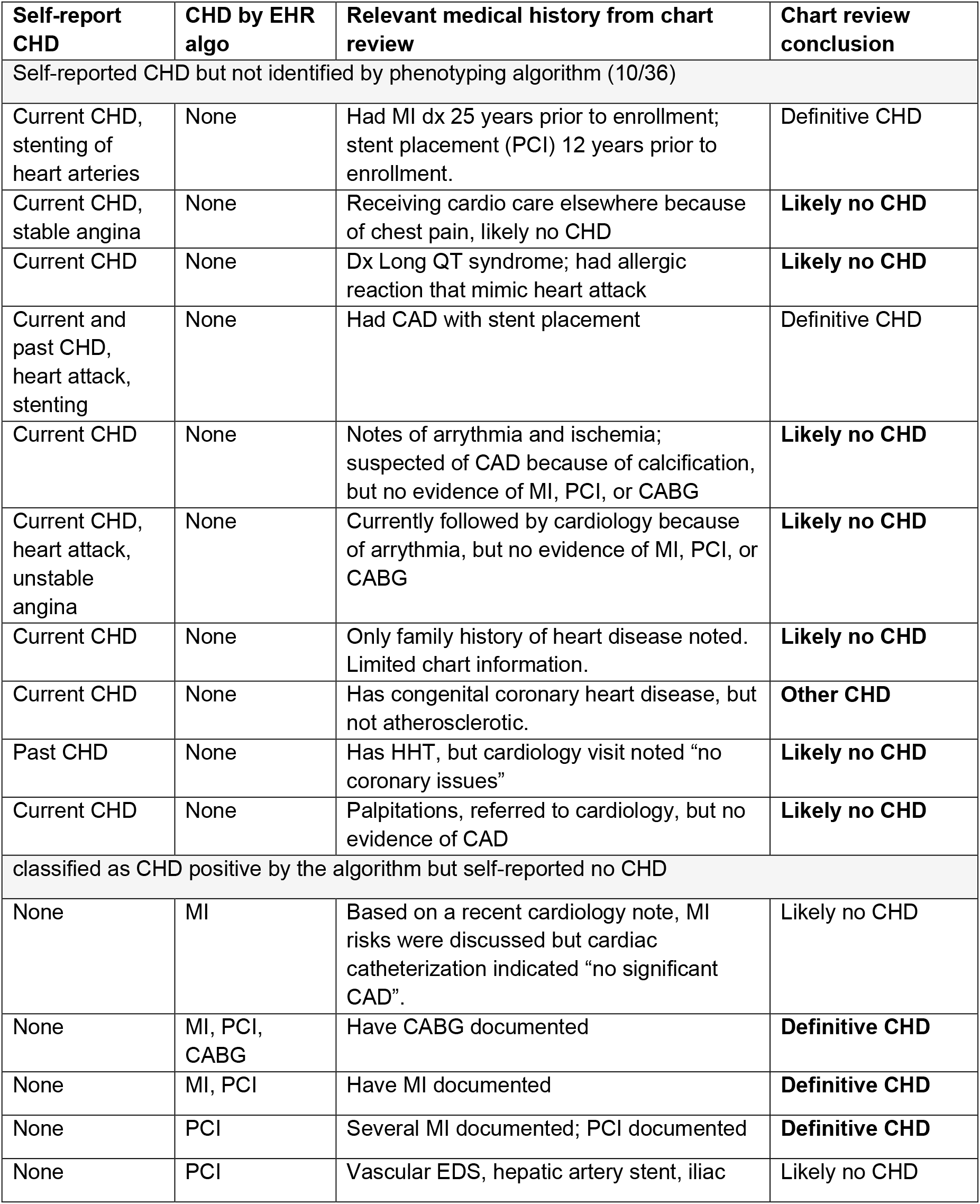
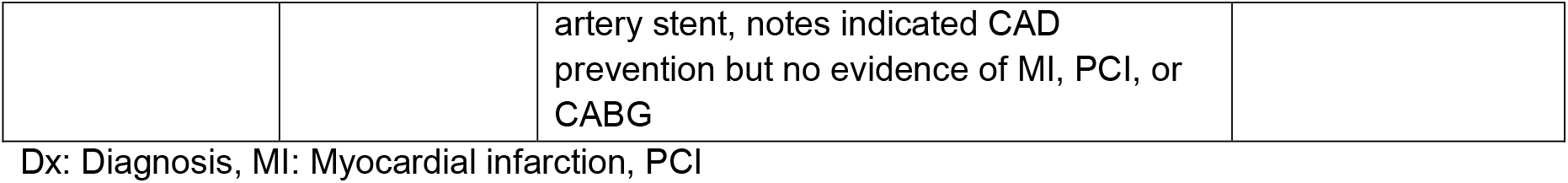
Chart review notes and conclusions for 15 participants with discordant CHD.

| **Self-report CHD** | **CHD by EHR algo** | **Relevant medical history from chart review** | **Chart review conclusion** |
| --- | --- | --- | --- |
| Self-reported CHD but not identified by phenotyping algorithm (10/36) |  |  |  |
| Current CHD, stenting of heart arteries | None | Had MI dx 25 years prior to enrollment; stent placement (PCI) 12 years prior to enrollment. | Definitive CHD |
| Current CHD, stable angina | None | Receiving cardio care elsewhere because of chest pain, likely no CHD | **Likely no CHD** |
| Current CHD | None | Dx Long QT syndrome; had allergic reaction that mimic heart attack | **Likely no CHD** |
| Current and past CHD, heart attack, stenting | None | Had CAD with stent placement | Definitive CHD |
| Current CHD | None | Notes of arrhythmia and ischemia; suspected of CAD because of calcification, but no evidence of MI, PCI, or CABG | **Likely no CHD** |
| Current CHD, heart attack, unstable angina | None | Currently followed by cardiology because of arrhythmia, but no evidence of MI, PCI, or CABG | **Likely no CHD** |
| Current CHD | None | Only family history of heart disease noted. Limited chart information. | **Likely no CHD** |
| Current CHD | None | Has congenital coronary heart disease, but not atherosclerotic. | **Other CHD** |
| Past CHD | None | Has HHT, but cardiology visit noted “no coronary issues” | **Likely no CHD** |
| Current CHD | None | Palpitations, referred to cardiology, but no evidence of CAD | **Likely no CHD** |
| classified as CHD positive by the algorithm but self-reported no CHD |  |  |  |
| None | MI | Based on a recent cardiology note, MI risks were discussed but cardiac catheterization indicated “no significant CAD”. | Likely no CHD |
| None | MI, PCI, CABG | Have CABG documented | **Definitive CHD** |
| None | MI, PCI | Have MI documented | **Definitive CHD** |
| None | PCI | Several MI documented; PCI documented | **Definitive CHD** |
| None | PCI | Vascular EDS, hepatic artery stent, iliac artery stent, notes indicated CAD prevention but no evidence of MI, PCI, or CABG | Likely no CHD |

Dx: Diagnosis, MI: Myocardial infarction, PCI

**Supplemental Table 4.** Cumulative incidence estimates [95% CI], under three alternative left truncation scenarios.

|  | Left truncation scenarios |  |  |
| --- | --- | --- | --- |
| Age (years) | First billing code as entry | Start of most recent consecutive period as entry | No left truncation |
| 40 | 0.008 [0.005, 0.015] | 0.015 [0.006, 0.037] | 0.001 [0.001, 0.002] |
| 45 | 0.016 [0.011, 0.024] | 0.027 [0.016, 0.046] | 0.004 [0.003, 0.006] |
| 50 | 0.025 [0.019, 0.032] | 0.039 [0.027, 0.057] | 0.008 [0.006, 0.010] |
| 55 | 0.043 [0.035, 0.051] | 0.063 [0.049, 0.080] | 0.018 [0.015, 0.021] |
| 60 | 0.072 [0.063, 0.082] | 0.099 [0.083, 0.117] | 0.038 [0.033, 0.043] |
| 65 | 0.110 [0.099, 0.122] | 0.143 [0.127, 0.162] | 0.070 [0.063, 0.078] |
| 70 | 0.141 [0.129, 0.155] | 0.179 [0.161, 0.198] | 0.101 [0.091, 0.111] |
| 75 | 0.188 [0.168, 0.209] | 0.225 [0.202, 0.249] | 0.149 [0.131, 0.169] |

## REFERENCE

1. Casey JA, Schwartz BS, Stewart WF, Adler NE. Using Electronic Health Records for Population Health Research: A Review of Methods and Applications. Annual Review of Public Health. 2016;37(Volume 37, 2016):61–81. doi:10.1146/annurev-publhealth-032315-021353

2. Bycroft C, Freeman C, Petkova D, et al. The UK Biobank resource with deep phenotyping and genomic data. Nature. 2018;562(7726):203–209. doi:10.1038/s41586-018-0579-z

3. The All of Us Research Program Investigators. The “All of Us” Research Program. New England Journal of Medicine. 2019;318(7):668–676.

4. Gianfrancesco MA, Goldstein ND. A narrative review on the validity of electronic health record-based research in epidemiology. BMC Medical Research Methodology. 2021;21(1):234. doi:10.1186/s12874-021-01416-5

5. Wei WQ, Denny JC. Extracting research-quality phenotypes from electronic health records to support precision medicine. Genome Med. 2015;7(1):41. doi:10.1186/s13073-015-0166-y

6. Wei WQ, Leibson CL, Ransom JE, et al. Impact of data fragmentation across healthcare centers on the accuracy of a high-throughput clinical phenotyping algorithm for specifying subjects with type 2 diabetes mellitus. Journal of the American Medical Informatics Association. 2012;19(2):219–224. doi:10.1136/amiajnl-2011-000597

7. O’Malley KJ, Cook KF, Price MD, Wildes KR, Hurdle JF, Ashton CM. Measuring diagnoses: ICD code accuracy. Health Serv Res. 2005;40(5 Pt 2):1620–1639. doi:10.1111/j.1475-6773.2005.00444.x

8. Patient-Centered Outcomes Research Institute. PCORI Methodology Standards. Patient-Centered Outcomes Research Institute; 2024.

9. Sulieman L, Cronin RM, Carroll RJ, et al. Comparing medical history data derived from electronic health records and survey answers in the All of Us Research Program. Journal of the American Medical Informatics Association. 2022;29(7):1131–1141. doi:10.1093/jamia/ocac046

10. Zozus MN, Walden A, Pieper CF. Comparing the Accuracy of Health Record Data and Self-Reported Data. Patient-Centered Outcomes Research Institute (PCORI); 2023. doi:10.25302/03.2023.ME.140922573

11. Weiskopf NG, Cohen AM, Hannan J, Jarmon T, Dorr DA. Towards augmenting structured EHR data: a comparison of manual chart review and patient self-report. AMIA Annu Symp Proc. 2020;2019:903–912.

12. Brown S, Lavery JA, Shen R, et al. Implications of Selection Bias Due to Delayed Study Entry in Clinical Genomic Studies. JAMA Oncol. 2022;8(2):287–291. doi:10.1001/jamaoncol.2021.5153

13. Betensky RA, Mandel M. Recognizing the problem of delayed entry in time-to-event studies: Better late than never for clinical neuroscientists. Annals of Neurology. 2015;78(6):839–844. doi:10.1002/ana.24538

14. Williams BA. Constructing Epidemiologic Cohorts from Electronic Health Record Data. Int J Environ Res Public Health. 2021;18(24):13193. doi:10.3390/ijerph182413193

15. Linder JE, Allworth A, Bland HT, et al. Returning integrated genomic risk and clinical recommendations: The eMERGE study. Genetics in Medicine. 2023;25(4). doi:10.1016/j.gim.2023.100006

16. Khera AV, Chaffin M, Aragam KG, et al. Genome-wide polygenic scores for common diseases identify individuals with risk equivalent to monogenic mutations. Nat Genet. 2018;50(9):1219–1224. doi:10.1038/s41588-018-0183-z

17. Dikilitas O, Schaid DJ, Kosel ML, et al. Predictive Utility of Polygenic Risk Scores for Coronary Heart Disease in Three Major Racial and Ethnic Groups. The American Journal of Human Genetics. 2020;106(5):707–716. doi:10.1016/j.ajhg.2020.04.002

18. Fahed AC, Wang M, Homburger JR, et al. Polygenic background modifies penetrance of monogenic variants for tier 1 genomic conditions. Nat Commun. 2020;11(1):3635. doi:10.1038/s41467-020-17374-3

19. Saadatagah S, Naderian M, Dikilitas O, Hamed ME, Bangash H, Kullo IJ. Polygenic Risk, Rare Variants, and Family History. JACC: Advances. 2023;2(7):100567. doi:10.1016/j.jacadv.2023.100567

20. Mosley JD, Gupta DK, Tan J, et al. Predictive Accuracy of a Polygenic Risk Score Compared With a Clinical Risk Score for Incident Coronary Heart Disease. JAMA. 2020;323(7):627–635. doi:10.1001/jama.2019.21782

21. Zhang Y, Dron JS, Bellows BK, et al. Familial Hypercholesterolemia Variant and Cardiovascular Risk in Individuals With Elevated Cholesterol. JAMA Cardiology. 2024;9(3):263–271. doi:10.1001/jamacardio.2023.5366

22. Limdi N, Beasley TM, Cortopassi J, et al. The Electronic Medical Records and Genomics study: Design and analytic framework for assessing the impact of genome-informed risk assessments. The American Journal of Human Genetics. Published online March 23, 2026. doi:10.1016/j.ajhg.2026.02.018

23. Lawson LP, Prows CA, Cortopassi J, et al. Return of genome-informed risk-assessment results for common conditions to 23,840 adults and children: An eMERGE network study. The American Journal of Human Genetics. 2026;113(4):678–691. doi:10.1016/j.ajhg.2026.02.016

24. Safarova MS, Liu H, Kullo IJ. Rapid identification of familial hypercholesterolemia from electronic health records: The SEARCH study. Journal of Clinical Lipidology. 2016;10(5):1230–1239. doi:10.1016/j.jacl.2016.08.001

25. Philofsky M, EHR Working Group. Observation Period Considerations for EHR Data. OMOP Common Data Model. Accessed July 9, 2025. https://ohdsi.github.io/CommonDataModel/ehrObsPeriods.html

26. Nykamp K, Anderson M, Powers M, et al. Sherloc: a comprehensive refinement of the ACMG–AMP variant classification criteria. Genetics in Medicine. 2017;19(10):1105–1117. doi:10.1038/gim.2017.37

27. Smith JL, Tcheandjieu C, Dikilitas O, et al. Multi-Ancestry Polygenic Risk Score for Coronary Heart Disease Based on an Ancestrally Diverse Genome-Wide Association Study and Population-Specific Optimization. Circ: Genomic and Precision Medicine. 2024;17(3). doi:10.1161/CIRCGEN.123.004272

28. Tcheandjieu C, Zhu X, Hilliard AT, et al. Large-scale genome-wide association study of coronary artery disease in genetically diverse populations. Nat Med. 2022;28(8):1679–1692. doi:10.1038/s41591-022-01891-3

29. Lennon NJ, Kottyan LC, Kachulis C, et al. Selection, optimization and validation of ten chronic disease polygenic risk scores for clinical implementation in diverse US populations. Nat Med. 2024;30(2):480–487. doi:10.1038/s41591-024-02796-z

30. Zou G. A Modified Poisson Regression Approach to Prospective Studies with Binary Data. American Journal of Epidemiology. 2004;159(7):702–706. doi:10.1093/aje/kwh090

31. Hosmer DW, Lemeshow S, May S. Applied survival analysis: regression modeling of time-to-event data. EBL. Published online 2008. doi:10.1002/9780470258019

32. Therneau TM. A Package for Survival Analysis in R. Published online 2024.

33. Kassambara A, Kosinski M, Biecek P. survminer: Drawing Survival Curves using “ggplot2.” Published online 2024. https://rpkgs.datanovia.com/survminer/index.html

34. Baierl J, Hsiao YW, Jones MR, Peng PC, Pharoah PDP. Measuring the accuracy of electronic health record-based phenotyping in the All of Us Research Program to optimize statistical power for genetic association testing. J Am Med Inform Assoc. Published online January 13, 2026:ocaf234. doi:10.1093/jamia/ocaf234

35. Korn EL, Graubard BI, Midthune D. Time-to-Event Analysis of Longitudinal Follow-up of a Survey: Choice of the Time-scale. American Journal of Epidemiology. 1997;145(1):72–80. doi:10.1093/oxfordjournals.aje.a009034

36. Irlmeier R, Hughey JJ, Bastarache L, Denny JC, Chen Q. Cox regression is robust to inaccurate EHR-extracted event time: an application to EHR-based GWAS. Bioinformatics. 2022;38(8):2297–2306. doi:10.1093/bioinformatics/btac086

37. Joseph A, Mullett C, Lilly C, et al. Coronary Artery Disease Phenotype Detection in an Academic Hospital System Setting. Appl Clin Inform. 2021;12(01):010–016. doi:10.1055/s-0040-1721012

38. Aragam KG, Dobbyn A, Judy R, et al. Limitations of Contemporary Guidelines for Managing Patients at High Genetic Risk of Coronary Artery Disease. Journal of the American College of Cardiology. 2020;75(22):2769–2780. doi:10.1016/j.jacc.2020.04.027

